# Prognostic accuracy of shock index for severe postpartum hemorrhage in high-income country: Protocol for a systematic review and meta-analysis

**DOI:** 10.1101/2021.05.17.21257068

**Authors:** Yuto Makino, Kentaro Miyake, Asami Okada, Yumie Ikeda, Yohei Okada

## Abstract

**Background:** This review aims to conduct a systematic review and meta-analysis to assess the prognostic value of shock index for prediction of severe Postpartum hemorrhage (PPH) in high-income countries.

**Method:** We will perform a systematic review and meta-analysis for diagnostic test accuracy (DTA). We will search CENTRAL, MEDLINE (Ovid), Web of Science, and other sources and include all relevant reports on the prognostic accuracy of shock index for severe PPH. The target condition is defined as severe PPH required higher-level care. Two review authors will independently screen the study eligibility and extract data from included studies. Two authors will also assess study quality using the QUADAS-2 (Quality Assessment of Diagnostic Accuracy Studies 2) tool. We will use the hierarchical models comprising both the bivariate model and the hierarchical summary receiver operating characteristic (HSROC) model for data synthesis if appropriate. We presented uncertainty of the accuracy estimates using 95% confidence intervals (CIs).

**Trial registration:** This review is submitted with University hospital medical information network clinical trial registry (UMIN-CTR) [UMIN 000044230].

**Conflicts of interest:** All authors declare to have no conflicts of interest

**Funding:** None

## Background

Deaths from obstetric hemorrhage, many of which are Postpartum hemorrhage (PPH), are preventable. Improving mortality is a global challenge.^1^ PPH is still one of the leading causes of maternal death.^2^ Severe PPH often requires multidisciplinary treatment besides the obstetric procedure.^3^ So, early detection of patients with severe PPH is important.^4^

Shock index can be easily calculated and has been used as a tool to predict critical bleeding in trauma patients, for example.^5^ Several studies in obstetrics also reported that shock index was helpful in predicting massive blood loss or the need for massive blood transfusion.^6 7 8^ However, the sensitivity and specificity were different in those studies. Previous studies, including systematic reviews, have not adequately assessed the prognostic accuracy of shock index for PPH. ^9^

Maternal mortality and the proportion of PPH differ between high-income countries and low-and middle-income countries.^2^ In high-income countries, rapid and accurate recognition of PPH can provide early decisions on higher-level care and more efficient use of health care resources. We aim to assess the prognostic value of shock index for prediction of severe PPH requiring higher-level care in high-income countries, through a systematic review and meta-analysis.

## Method

We will perform a systematic review and meta-analysis for diagnostic test accuracy (DTA). This review will be conducted according to the Preferred Reporting Items for a Systematic Review and Meta-analysis of Diagnostic Test Accuracy Studies (The PRISMA-DTA Statement)^10^ and the Cochrane Handbook for Systematic Reviews of Diagnostic Test Accuracy.^11^ We submitted the review protocol to the pre-print server (medRxiv) and registered with University hospital medical information network (UMIN) clinical trial registry [UMIN 000044230].

### Criteria for considering studies for this review

#### Types of studies

We will include prospective or retrospective cohort studies and cross-sectional studies evaluating the prognostic accuracy of shock index for PPH, with enough details to calculate study - specific sensitivity and specificity. We will exclude studies reporting insufficient data for constructing a two- by-two table. We will also exclude case reports and diagnostic case-control studies.

#### Participants

We will include women in the peripartum period. We will exclude pregnant women assessed only prepartum period.

#### Setting

We will include the studies in high-income countries defined by World Bank Country and Lending Groups^12^ regardless of the types of the medical institution (e.g., primary care setting or tertiary hospital).

#### Index test

Index test is shock index defined as heart rate divided by systolic blood pressure. Any cutoffs will be included. We will only include the measurement timing was during primary assessment (i.e., before higher-level care). Other indexes such as blood pressure, heart rate, etc. evaluated in the included studies will be extracted and assessed.

#### Target conditions

We will include studies evaluating the incidence of severe PPH or the adverse events associated with PPH. We will assess severe PPH required higher-level care as a primary outcome for a meta-analysis. It is defined as the need for use of blood products, interventional radiology, laparotomy (other than cesarean section), or admission to intensive care unit (ICU). This definition is based on WHO “Interventions based” criteria to identify maternal near miss.^13^ We will also assess a blood loss of 1000 ml or more within 24 hours after birth as a secondary outcome for a meta-analysis. This is a commonly used definition of severe PPH.^14^

### Search methods for identification of studies

#### Electronic searches

We have developed a comprehensive search strategy in collaboration with Kyoto University Medical Librarian. We will search the following databases for relevant reports regardless of language or publication status (published, unpublished, in press, or in progress):

- The Cochrane Central Register of Controlled Trials (CENTRAL), in the Cochrane Library
- MEDLINE (Ovid SP, 1946 to present)
- Web of Science (1945 to present)

The search strategy for MEDLINE can be found in Appendix 1. We will also conduct a search of ClinicalTrials.gov (clinicaltrials.gov/) and the World Health Organization International Clinical Trial Registry Platform (WHO ICTPR) Search Portal (apps.who.int/trialsearch/) for ongoing or unpublished trials.

#### Searching other resources

We will check the reference lists of review articles and scan recent meta-analysis or reviews on the topic. We will also examine conference abstracts of the society related to the topic. If necessary, we will contact trial authors for additional information.

### Data collection and analysis

#### Selection of studies

We will use the search strategy described above to obtain titles and abstracts of the studies. Two review authors will independently screen titles and abstracts of the results of the search to identify potentially relevant studies. We will obtain the full text of these studies and exclude studies that do not meet the eligibility. We will provide a list of excluded studies with reasons for exclusion. We will resolve conflicts by discussion. If we are unable to reach a consensus, we will consult the third review author.

### Data extraction and management

We will use a data collection form for extracting the following study information:

- Methods: study design, total duration of the study, number of study centers and country, study setting, and date of the study.
- Participants: number of participants, age, body mass index (BMI), cause of PPH (e.g., atonic bleeding or placenta previa), other complications (e.g., infection, cardiomyopathy), type of anesthesia, inclusion criteria, and exclusion criteria.
- Index test: timing of shock index measurement, cutoff for shock index, indexes other than shock index (e.g., blood pressure, heart rate, lactate).
- Target conditions: definition of severe PPH given in each study.
- Outcomes: From the 2 × 2 tables, we will calculate true positives, false positives, true negatives, and false negatives.
- Notes: funding for trial, and notable conflicts of interest of authors.

Two review authors will independently extract the data using standard data extraction forms. Studies reported in journals in languages other than English will be translated prior to assessment. Where multiple publications of a single study exist, we will group the reports and extract the most likely correct data from multiple reports for analysis. We will resolve conflicts on data extraction by discussion. If we are unable to reach a consensus, we will consult the third review author. If additional information is needed, one review author will contact the corresponding author of the study. After we have completed data extraction, another review author will check the data.

### Assessment of methodological quality

Two review authors will independently assess the methodological quality of the included studies using the QUADAS-2 tool.^15^ We will tailor the QUADAS-2 tool to this review. QUADAS-2 consists of four domains: patient selection, index test, reference standard, and flow and timing. We will assess all four domains for potential risk of bias and the first three domains for concerns of applicability. We will resolve conflicts by discussion. If we are unable to reach a consensus, we will consult the third review author. We will present the results of the quality assessment in tables, graphs, and in the text.

### Statistical analysis and data synthesis

We will present the results of individual studies by plotting sensitivity, specificity, and 95% confidence interval extracted from each study on 1-dimensional forest plots (ordered by specificity) and on the receiver-operating characteristic (ROC) space. We will use the hierarchical (mixed) models comprising both the bivariate model and the hierarchical summary ROC (HSROC) model for data synthesis.^16 17^ The HSROC analysis would be necessary where we have a diagnostic tool that provides a result on a continuous scale, where the cut-off point varies between studies. We will only conduct HSROC analysis if appropriate. We will fix specificity at the median value of included studies. We will present a summary estimate of the sensitivity and specificity of clinically commonly used shock index cutoffs in bivariate mode as a secondary analysis. We will report data narratively if it is not appropriate to combine in a meta-analysis.

We will perform the same analysis for indexes other than shock index as additional analysis, if possible. All analyses will be performed using Review Manager 5.3 (Cochrane Collaboration, London, United Kingdom) and commonly available statistical software such as STATA (version 16. College Station, TX USA: StataCorp, 2019).

### Investigations of heterogeneity

We will visually inspect forest plots and summary ROC curve to assess heterogeneity between studies. We plan to carry out the following subgroup analyses if we find sufficient data from the included studies:

- Setting (e.g., primary care setting versus tertiary hospital)
- Mode of delivery: vaginal versus cesarean section
- Immediately after transportation versus during hospitalization
- Cause of PPH (e.g., atonic bleeding or placenta previa)
- Definition of severe PPH within the included studies (e.g., need for use of blood products only or; need for surgical hemostasis only
- Presence of other complications (e.g., infection or pulmonary embolism)
- Timing of shock index measurement
- Type of anesthesia: regional versus general

### Sensitivity analyses

We plan to carry out the following sensitivity analyses, to test whether key methodological factors or decisions have affected the main result:

- Including studies only assessed low risk of bias
- The different definition of severe PPH (e.g., need for transfusion of more than 10 units of blood, need for laparotomy)

### Assessment of reporting bias

We will not perform a quantitative assessment of reporting bias because there is a lack of consensus about the most robust approach to assessment of reporting bias in DTA.^18^ We will, however, try to minimize the risk of publication bias by using a robust and comprehensive search strategy.

## Data Availability

Not applicable

## Appendix 1

Database: Ovid MEDLINE(R) ALL <1946 to Present>

Search Strategy:

--------------------------------------------------------------------------------

1. “Shock index”.mp.
2. exp Blood Pressure/
3. “Blood pressure”.ti,ab,kf.
4. exp Heart Rate/
5. (Heart rate or Heart rates or Cardiac Rate or Cardiac Rates or Pulse Rate or Pulse Rates or Cardiac Chronotropy or Cardiac Chronotropism).ti,ab,kf.
6. or/1-5
7. exp Postpartum Hemorrhage/
8. exp Uterine Hemorrhage/
9. (Uterine Hemorrhage or Uterine Hemorrhages or Uterine Bleeding or Uterine Bleedings or Vaginal Bleeding or Vaginal Bleedings).ti,ab,kf.
10. ((post partum or postpartum or postpartal or postnatal or post natal or puerperal or after childbirth or after birth or after giving birth or after delivery or after the delivery or following childbirth) adj3 (haemorrhag* or hemorrhag* or bleed* or bloodloss* or blood loss* or coagulopath*)).ti,ab,kf.
11. or/7-10
12. exp animals/ not humans.sh.
13. (6 and 11) not 12

